# Infratentorial White Matter Integrity as a Potential Biomarker for Post-Stroke Aphasia

**DOI:** 10.1101/2024.11.21.24317584

**Authors:** Ben Zhang, Caroline Schnakers, Kevin Xing-Long Wang, Jing Wang, Sharon Lee, Henry Millan, Melissa Howard, Emily Rosario, Zhong Sheng Zheng

## Abstract

Traditionally, neuroimaging studies of post-stroke aphasia focus on supratentorial brain regions related to language function and recovery. However, stroke-induced lesions often distort these areas, posing a challenge for neuroimaging analyses aimed at identifying reliable biomarkers. This study seeks to explore alternative biomarkers in regions less affected by direct stroke damage, such as white matter regions below the tentorium, to overcome these methodological limitations. Diffusion tensor imaging was accomplished on 55 participants with chronic post-stroke aphasia. Focusing on regions below the tentorium, correlations were analyzed between Western Aphasia Battery-Revised scores and average fractional anisotropy values. The volume of intersection between each participant’s lesion and their left arcuate fasciculus was also analyzed for correlations with Western Aphasia Battery-Revised scores as well. Linear regression analyses were then conducted using regions showing significant correlations as univariate predictors. After applying multiple comparisons corrections, we found that average fractional anisotropy in the middle cerebellar peduncle was positively correlated with aphasia quotient (p = 0.004), spontaneous speech (p = 0.005), auditory verbal comprehension (p = 0.004), naming & word finding (p = 0.005) and repetition (p = 0.013). Average fractional anisotropy in the left inferior cerebellar peduncle positively correlated with spontaneous speech (p = 0.018) and auditory verbal comprehension (p = 0.018). Average fractional anisotropy in the left corticospinal tract positively correlated with aphasia quotient (p = 0.019) and spontaneous speech (p = 0.005). Volume of intersection between the left arcuate fasciculus and participant lesion was negatively correlated with aphasia quotient (p = 0.014) and repetition (p = 0.002). Through linear regression analyses, average fractional anisotropy of the middle cerebellar peduncle significantly predicted aphasia quotient and all sub-scores. Average fractional anisotropy of the left inferior cerebellar peduncle significantly predicted all scores except repetition. Average fractional anisotropy of the left corticospinal tract significantly predicted all scores except for auditory verbal comprehension. Volume of intersection between left arcuate fasciculus and lesions significantly predicted all scores except for auditory verbal comprehension. These findings underscore the potential of infratentorial white matter regions as biomarkers of aphasia severity, encompassing overall and specific subdomain impairment. By shifting the focus to below the tentorium, it becomes possible to find more robust targets for further research and therapeutic interventions. This approach is not only able to sidestep analytical complications posed by cortical lesions, it also opens new doors for understanding complex cerebellar mechanisms that underlie language function and recovery post stroke.

## INTRODUCTION

Aphasia is a complex neurological condition that affects millions globally, characterized by deficits in language production and comprehension commonly resulting from stroke.^1^ Research in post-stroke aphasia (PSA) has primarily focused on supratentorial regions, including cortical language areas (e.g., Broca’s and Wernicke’s areas) and associated white matter tracts (e.g., arcuate fasciculus).^1-5^ However, when dealing with brain distorting lesions resulting from supratentorial stroke, this limited perspective leads to methodological challenges in research and clinical assessments. While exploring cortical contributions to aphasia has provided valuable insights, the significance of white matter integrity below the tentorium (infratentorium) and its relationship to PSA has often been overlooked^3-4^

Language processing and speech production rely on an extensive network that exceeds the well-known Broca’s and Wernicke’s areas, engaging a myriad of cortical, subcortical, and cerebellar regions.^6-10^ Recent research has demonstrated the importance of the cerebro-cerebellar network, with the cerebellum—situated beneath the tentorium—playing a crucial role.^11^ The cerebellum is consistently implicated in language function, playing a role in motor components such as articulation, as well as nonmotor aspects of language such as semantics.^12^ To touch on motor components, neuroimaging studies have shown activation of representations of motor effectors in the cerebellum during tongue movement or articulation.^11-12^ Furthermore, early clinical evidence of perceptual deficits in temporal discrimination tasks associated with cerebellar pathology show how the cerebellum partially functions as an “internal clock” for motor and perceptual timing. Consistent with this, dysfunction in infratentorial regions appears to impair temporal aspects of speech perception, compromising fluent speech.^8-9,11-13^. Non-motor aspects of language, such as phoneme identification and speech distortion adaptation, have also been shown to be correlated with the cerebellum, particularly the right Crus I.^11,13^ Many studies also report semantic deficits in patients with cerebellar dysfunction.^12^ Additionally, observed cognitive deficits such as verbal learning and working memory may have an anatomical basis in the reciprocal fiber systems connecting the cerebellum with the cerebral association cortex.^13^

Following a brain injury, white matter fibers may shift, causing disconnections that necessitate neural reorganization and adaptation.^14^ This reorganization process is critical for recovery from aphasia, as demonstrated by research linking changes in white matter integrity to improvements in language functions.^15^ The relevance of white matter pathways to speech challenges, including issues with repetition, comprehension, and naming, has also been increasingly recognized.^16-18^ Despite these insights, however, the role of infratentorial white matter integrity in PSA remains understudied. This gap in understanding is especially concerning due to the issues posed during analysis of supratentorial regions in PSA patients, as lesions often distort and complicate accurate delineation of white matter tracts.

Our study focuses on addressing this gap by exploring the association between white matter integrity below the tentorium and language function in patients with chronic PSA. By focusing on infratentorial regions, which are generally less affected in PSA, we aim to identify potential biomarkers that could lead to more accessible and clinically reliable measures of post-stroke aphasia.

Utilizing Diffusion Tensor Imaging (DTI), a pivotal method for evaluating white matter integrity, we investigated the microstructural properties of infratentorial white matter tracts alongside their relationship to language function as measured by the Western Aphasia Battery—Revised (WAB-R).^19-20^ As many DTI studies have been supratentorially focused, our approach opens up new avenues for understanding intricate changes in language-related white matter following stroke by circumventing challenges from cortical damage.^21-23^

The results of this study could have significant implications for both research and clinical practice in PSA. By investigating the relationships between white matter integrity in infratentorial brain regions and language function, we may help provide a more encompassing understanding of neural correlates of PSA, as well as potentially offer new and more robust targets for therapeutic interventions and prognostic indicators.

## MATERIALS AND METHODS

### Participants

The dataset (behavioral and diffusion-weighted data) used in this analysis was derived from the pre-intervention (i.e., baseline) data of two previous clinical trials that investigated the efficacy of transcranial direct current stimulation (t-DCS) in treating post-stroke aphasia. While the original trials included both pre- and post-intervention timepoints, only the pre-intervention data were utilized in the current study. This approach allowed for the combination of data from both trials, resulting in a larger dataset for analysis. The sample included 57 participants with chronic PSA, excluding two participants with right hemisphere-only lesions for a final cohort of 55. This final cohort consisted of predominantly left hemisphere strokes (n = 42), with a subset of participants presenting bilateral lesions (n = 13). The group lesion overlap of the patients is shown in Figure 1. The inclusion criteria included individuals with PSA (diagnosed by a Neurologist or Speech/Language Therapist) without a history of other neurological disorders, were at least 18 years or older, 6 months post-stroke, and English proficient. Clinical as well as demographic data is included in Table 1. This study was approved by the Institutional Review Board at Casa Colina Hospital and Centers for Healthcare, and all participants provided written informed consent prior to enrolling.

**Table 1.** Patient demographics and clinical information.

| Table 1. Patient Demographics and Clinical Information |  |
| --- | --- |
| Variable | N |
| Age (years) |  |
| Mean | 60.3 ± 3.67 |
| Time Since Injury (days) |  |
| Mean | 1440.3 ± 344.54 |
| Aphasia Quotient |  |
| Mean | 57.32 ± 7.23 |
| Gender |  |
| Male | 44 |
| Female | 11 |
| Stroke Type (Ischemic / Hemorrhagic / Both / Unsure) |  |
| Ischemic | 42 |
| Hemorrhagic | 10 |
| Both | 2 |
| Unsure | 1 |
| Fluency (Fluent / Non-Fluent) |  |
| Fluent | 25 |
| Non-Fluent | 30 |
| Lesion Type |  |
| Left Hemisphere | 42 |
| Bilateral |  |
| Bilateral Cortical | 5 |
| Bilateral Subcortical | 3 |
| Left Cortical + Right Subcortical | 5 |

**Figure 1.**
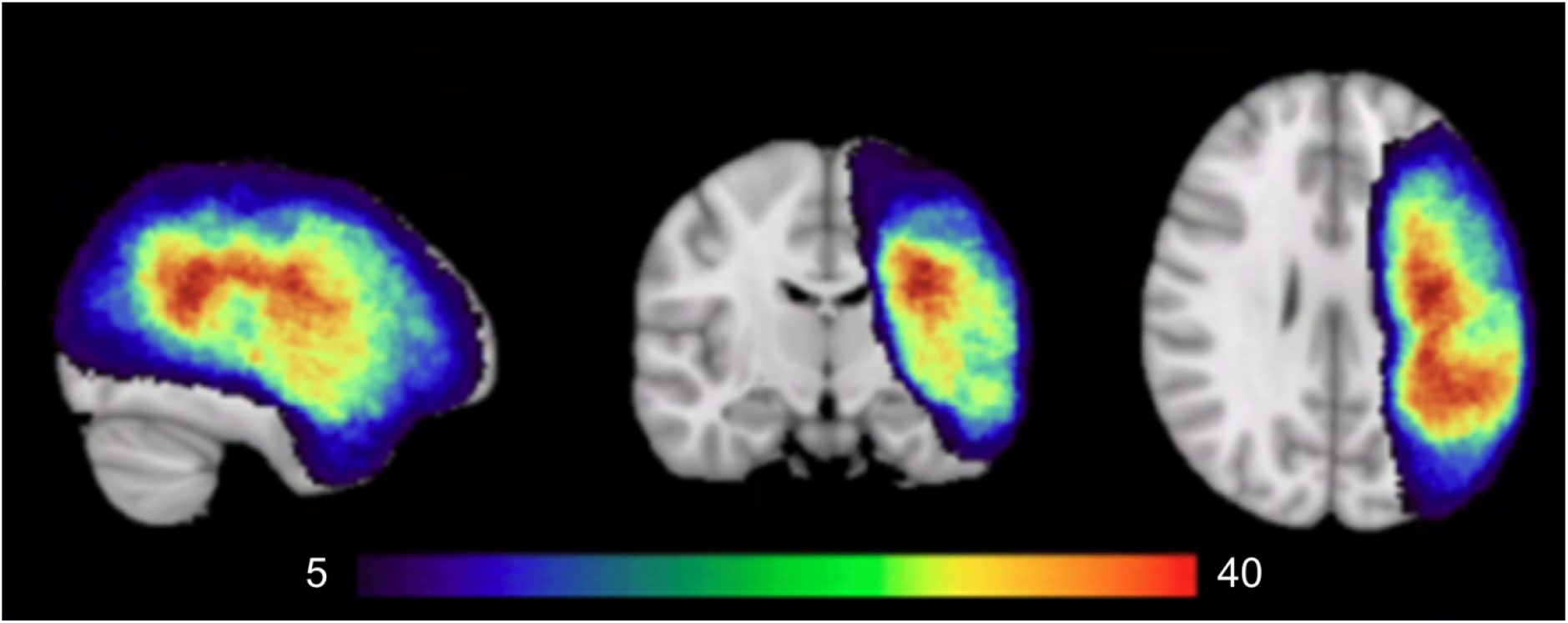
Group lesion overlap. The group lesion map indicates areas of stroke damage shared by at least five participants. Lesion overlap was most pronounced in the left perisylvian regions, particularly within the left superior longitudinal fasciculus. The color bar indicates the number of participants with overlapping lesions in each region.

### Language Assessment

All participants underwent aphasia assessment using the WAB-R. This comprehensive assessment evaluates language capabilities across four dimensions: spontaneous speech (SS), auditory verbal comprehension (AVC), naming and word finding (NW), and repetition (REP). Additionally, the WAB-R provides an overall aphasia quotient (AQ) [range 0-100], which encompasses the four subscores and provides a measure of aphasia severity, with higher scores indicating better language performance.

### MRI Acquisition

A 3T Siemens Magnetom Verio scanner was used to acquire MRI data. Using T1-weighted Magnetization Prepared Rapid Gradient Echo (MPRAGE) sequence (TR = 2300 ms; TE = 2 ms; flip angle = 9°; FOV = 230 mm; slice thickness = 1 mm with no gap; # of slices = 160; matrix size = 224 × 224), a high resolution 3D anatomical scan was collected. Diffusion-weighted imaging was then performed with following parameters : TR = 10900 ms; TE = 95 ms; slice thickness = 2 mm with no gap; # of slices = 82; matrix size = 122 × 122. Diffusion sensitizing gradients were applied along 64 non-collinear directions (b = 1000 s/mm^2^). Five images without no diffusion weighting (b = 0) were also obtained.

### Data Analysis

#### Preprocessing & Lesion Identification

Initially, T1-weighted images were subjected to bias-field correction utilizing FMRIB Software Library (FSL) tools (http://www.fmrib.ox.ac.uk/fsl). Following this, the Optimized Brain Extraction for Pathological Brains (optiBET) tool was employed for brain extraction.^2^ Stroke lesions were delineated using the automated segmentation tool LINDA (Lesion Identification with Neighborhood Data Analysis), which processes the patients’ T1-weighted images.^25^ Subsequently, lesion masks generated by LINDA were manually refined as necessary. For standard space alignment of individual T1-weighted lesion masks (MNI 1mm), we utilized ANTs (Advanced Normalization Tools).^26^ These standardized lesion masks were then binarized and aggregated to create a group lesion map, shown in Figure 1. The preprocessing of DWI data was conducted with DTIPrep, which served to identify and eliminate artifact-laden volumes, alongside corrections for eddy currents and head motion via affine registration to the average b=0 reference image. ^27^ The skull stripping of the b=0 image was accomplished using the Brain Extraction Tool (BET).^28^

### DTI Analysis

After preprocessing, diffusion tensors were calculated at each voxel of the brain using the FSL’s diffusion toolbox (FDT). From these tensors, fractional anisotropy (FA) maps were computed for each participant.

We employed a region of interest (ROI) approach using the John Hopkins University (JHU) white matter atlas (http://cmrm.med.jhmi.edu). Due to the extensiveness and heterogeneity of the lesions, which primarily affected regions above the tentorium, accurate delineation of supratentorial ROIs proved challenging. Therefore, we focused our investigation on infratentorial regions, which were predominantly intact across all patients. Twelve ROIs were selected (Figure 2): middle cerebellar peduncle (MCP), pontine crossing tract (PCT), bilateral medial lemniscus (ML), bilateral cerebral peduncle (CP), bilateral corticospinal tract (CST), bilateral superior cerebellar peduncle (SCP), and bilateral inferior cerebellar peduncle (ICP). Applying nonlinear registration between the standard space (FMRIB58_FA) and patients’ diffusion space, the standard ROIs were transformed into the latter. Before analysis of data, ROIs were examined visually for accuracy and corrected manually if required. A whole-brain white matter segmentation mask was generated for each patient to intersect with the ROIs, ensuring only white matter voxels are included. Average FA of each ROI was calculated for each patient, providing a measure of white matter integrity.

**Figure 2.**
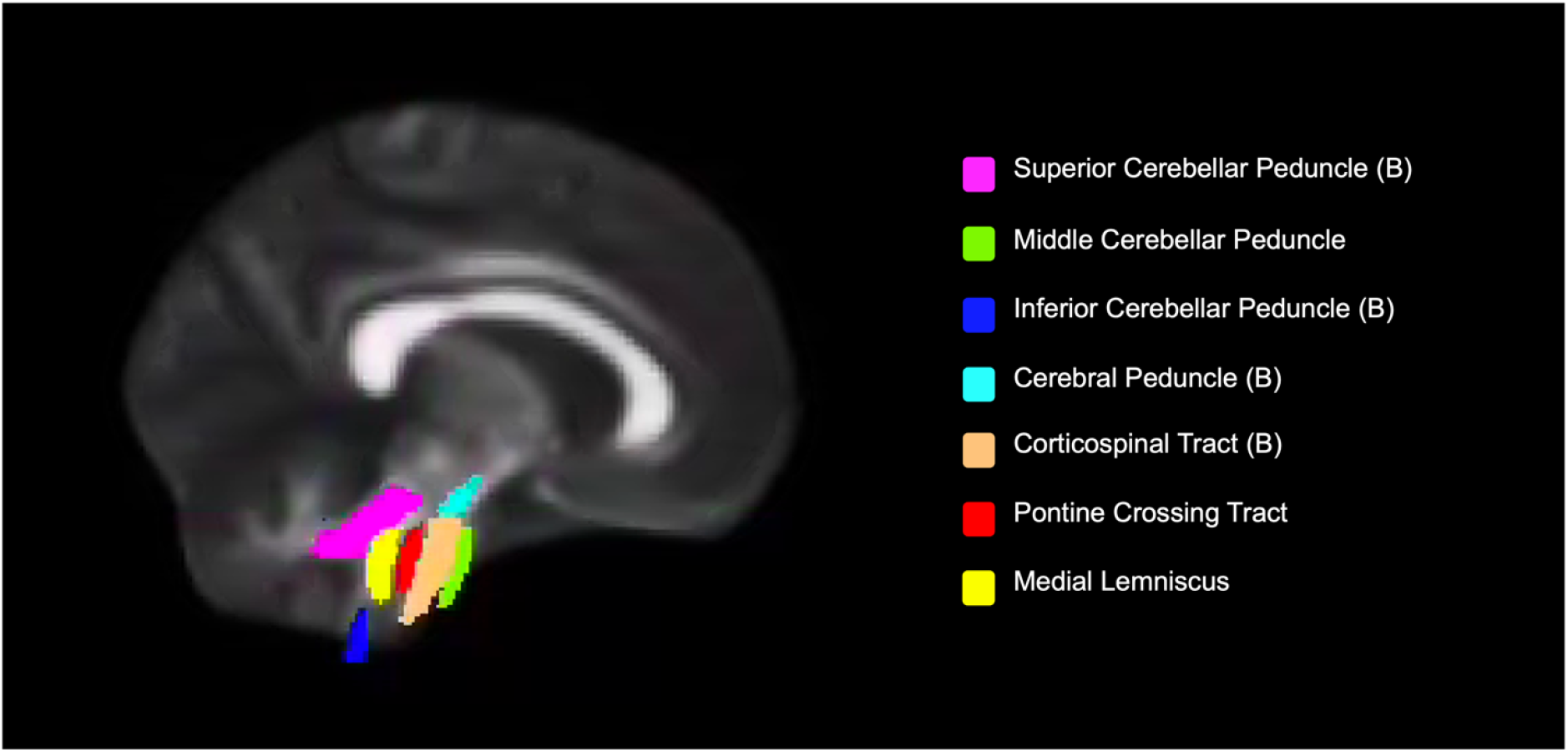
Infratentorial White Matter Regions of Interest. Sagittal view displaying infratentorial white matter labels from the Johns Hopkins White Matter Atlas. (B) = Bilateral structures

### Arcuate Lesion Calculation

Given the critical role of the arcuate fasciculus in language processing, the extent of damage to this tract likely has a significant impact on the severity of PSA.^2-5^ Therefore, we opted to calculate the overlap between each patient’s stroke lesion and their arcuate fasciculus to better understand its impact on their condition, as well as compare its effect on PSA against that of infratentorial white matter tracts.

A tractography based atlas of the arcuate fasciculus was obtained from NatBrainLab (https://www.natbrainlab.co.uk/atlas-maps). The FLIRT tool for linear registration was employed against the MNI152 standard brain template, transforming the atlas from its native space into the standardized MNI space. The transformed atlas was then binarized and separated into left and right segments to facilitate the subsequent overlap analysis. Each participant’s T1-weighted MRI scan was standardized to the MNI space and binarized as well.

Binarized lesion mask were then intersected with the binarized arcuate fasciculus ROI using the fslmaths command. This procedure generated new images for each subject, representing the overlap between the lesion and the left and right arcuate segments, thereby isolating the affected portions of the tract. The volume of overlap (arcuate lesion volume), indicative of the extent to which the arcuate fasciculus was compromised by the lesion, was quantified using the fslstats command.

### Statistical Analysis

To assess baseline differences among participants, we initially performed Spearman’s rank correlations between the mean FA of each ROI, WAB-R scores, age, and time since injury (TSI). Given that neither age nor TSI exhibited significant correlations with the variables of interest, no covariates were included in subsequent correlation analyses.

Spearman’s rank correlations were then conducted, exploring the relationship between baseline average FA in infratentorial regions and WAB-R scores after stroke. For comparative purposes, we also analyzed Spearman’s correlations between left arcuate lesion volume and WAB-R scores, as most participants had lesions affecting this canonical language structure. While white matter integrity measures like FA could be informative, especially in infratentorial regions (relatively spared in PSA), the extensive lesions in our cohort precluded reliable brain registration in regions above the tentorium. This limitation necessitated that our supratentorial focus be on lesion volume. The Benjamini-Hochberg method (FDR = 0.05) was utilized for multiple comparisons correction.

Building on these findings, univariate linear regression analyses were conducted to pinpoint the which significant white matter regions served as reliable predictors of WAB-R scores. A univariate linear regression analysis between the arcuate lesion volumes and WAB-R scores was carried out as well for the purpose of comparison.

## Results

The group lesion overlap map portrayed damage mostly focused in the left perisylvian regions, with the largest overlap occupying the left superior longitudinal fasciculus (see Figure 1), which has substantial anatomical overlap with the left arcuate fasciculus.

Following correction for multiple comparisons, significant correlations were observed between the mean FA in the middle cerebellar peduncle (MCP) and all WAB-R scores. Specifically, correlations were found with the AQ (r_s_ = 0.388, p = 0.004), SS (r_s_ = 0.383, p = 0.005), AVC (r_s_ = 0.391, p = 0.004), NW (r_s_ = 0.379, p = 0.005), and REP (r_s_ = 0.340, p = 0.013). Similarly, FA in the left inferior cerebellar peduncle (L-ICP) correlated with SS (r_s_ = 0.323, p = 0.018) and AVC (r_s_ = 0.323, p = 0.018). Additionally, FA in the left corticospinal tract (L-CST) correlated with AQ (r_s_ = 0.321, p = 0.019), and SS (r_s_ = 0.384, p = 0.005). Graphical representations of these correlations are presented in Figures 3-6. For the sake of comparison, we found that after multiple comparisons correction the left arcuate lesion volume correlated with WAB-R scores as well, specifically AQ (r_s_ = -0.352, p = 0.014) and REP (r_s_ = -0.433, p = 0.002).

**Figure 3.**
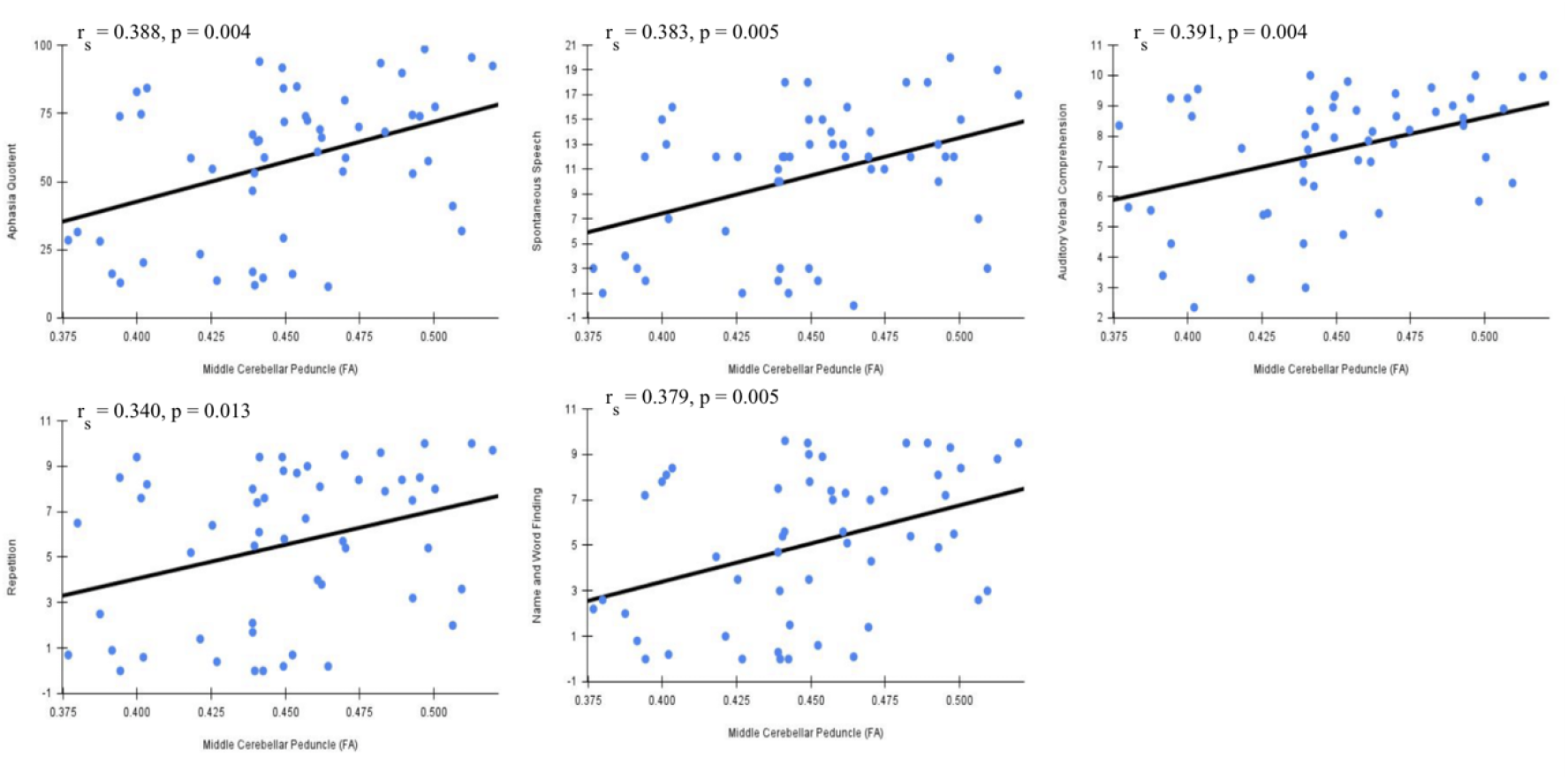
Middle Cerebellar Peduncle Correlations with WAB-R Scores. Scatterplots indicating the correlation between the average FA of MCP and WAB-R scores

**Figure 4.**
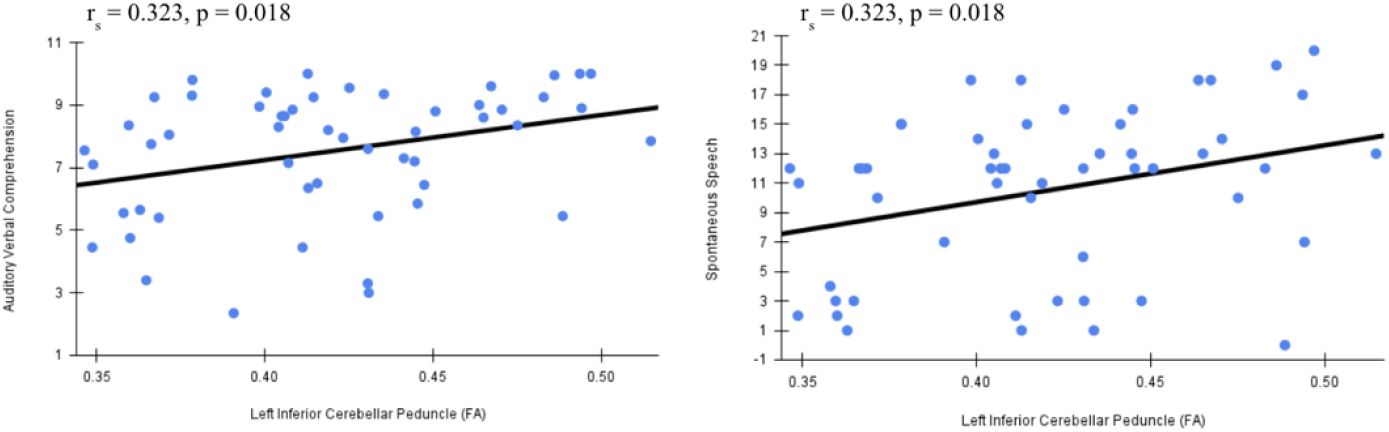
Left Inferior Cerebellar Peduncle Correlations with WAB-R Scores. Scatterplots indicating the correlation between the average FA of L-ICP and WAB-R scores

**Figure 5.**
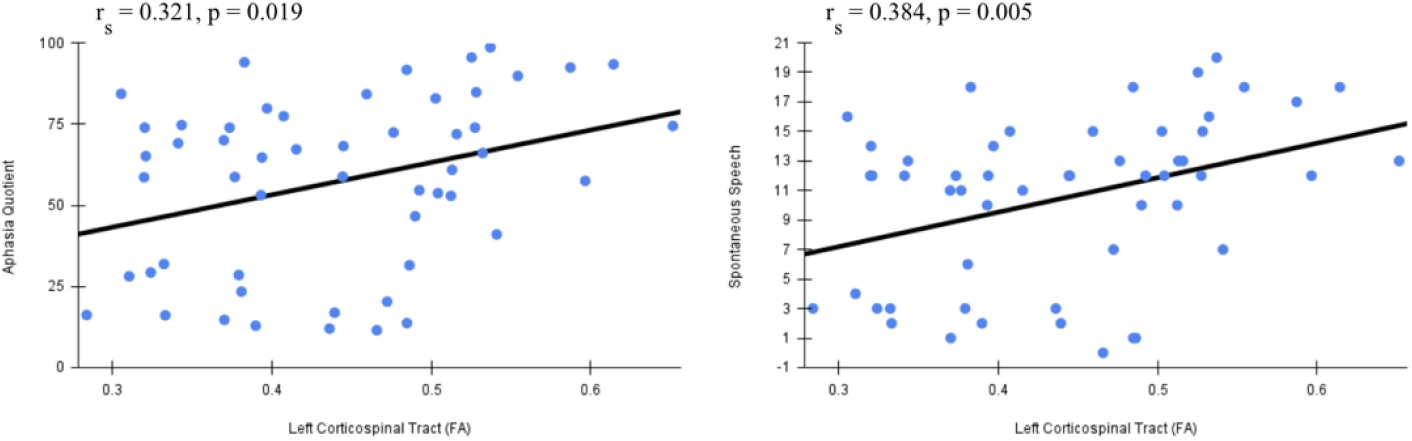
Left Corticospinal Tract Correlations with WAB-R Scores. Scatterplots indicating the correlation between average FA of L-CST and WAB-R scores

**Figure 6.**
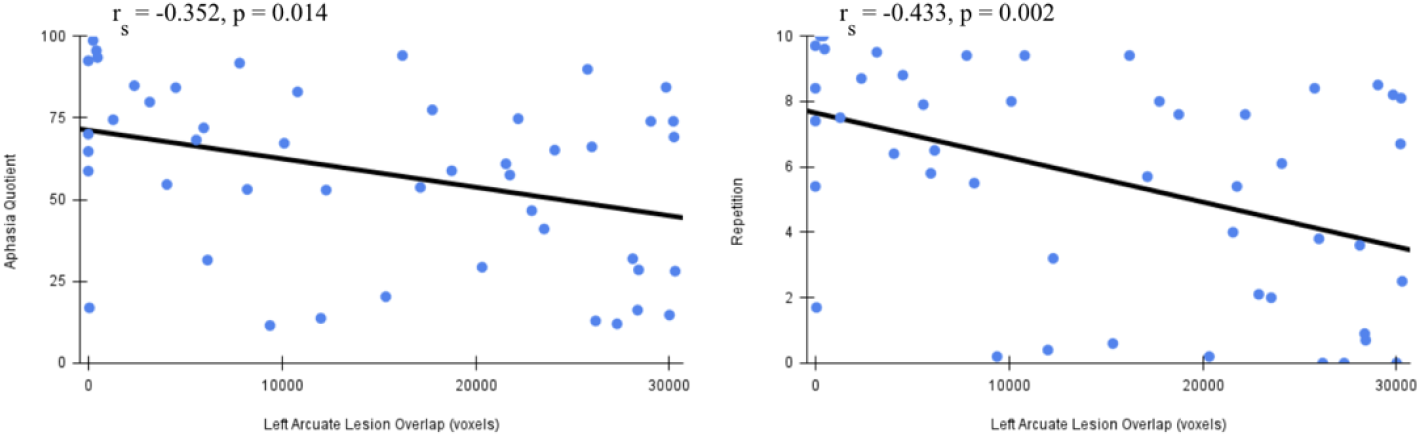
Left Arcuate Lesion Volume Correlations. Scatterplots indicating the correlation between Left Arcuate Lesion Volume and WAB-R scores

To assess the predictive value of FA in specific white matter tracts on aphasia severity, we followed our correlation analyses with univariate linear regression analyses. WAB-R scores served as dependent variables, while FA values in the MCP, L-CST, and L-ICP were used as independent variables. FA in the MCP emerged as a significant univariate predictor for all WAB-R scores: AQ (t(55) = 3.163, p = 0.003), SS (t(55) = 3.166, p = 0.003), AVC (t(55) = 3.117, p = 0.003), NW (t(55) = 2.946, p = 0.005), and REP (t(55) = 2.464, p = 0.017). FA in the L-ICP significantly predicted four out of five WAB-R scores (except for REP): AQ (t(55) = 2.145, p = 0.037), SS (t(55) = 2.338, p = 0.023), AVC (t(55) = 2.424, p = 0.019), and NW (t(55) = 2.263, p = 0.028). Similarly FA in the L-CST demonstrated significant predictive value for the following measures: AQ (t(55) = 2.543, p = 0.014), SS (t(55) = 2.907, p = 0.005), NW (t(55) = 2.193, p = 0.033), and REP (t(55) = 2.030, p = 0.048). The left arcuate lesion volume showed statistical significance as a predictor of AQ (t(55) = -2.599, p = 0.013), SS (t(55) = -2.198, p = 0.033), NW (t(55) = -2.171, p = 0.035), and REP (t(55) = -3.366, p = 0.002).

## DISCUSSION

Our study aimed to explore the potential significance of infratentorial white matter regions in post-stroke aphasia (PSA) by investigating the relationship between language performance and infratentorial white matter integrity. This approach expands upon traditional focuses on cortical language areas and supratentorial white matter tracts, offering a unique advantage in that it is less affected by PSA lesions, which predominantly occur in supratentorial areas. By examining regions that are typically preserved in PSA, we provide a more consistent basis for assessing language function across patients with varying lesion profiles. Our findings can be summarized as follows: 1) mean FA values of the middle cerebellar peduncle (MCP), left inferior cerebellar peduncle (L-ICP), left corticospinal tract (L-CST), and also left arcuate fasciculus lesion volume were positively correlated with Western Aphasia Battery-Revised (WAB-R) scores; 2) These regions demonstrated significance as univariate predictors of WAB-R scores in linear regression models.

Fractional anisotropy, a measurement for the directionality of white matter microstructures such as myelin, microtubules, and axons, acts as a proxy for white matter integrity.^29^ Lower levels of FA have been found to be associated with worse behavioral and motor outcomes following brain injury.^30-33^ Better baseline integrity of the MCP, L-ICP, and L-CST following PSA could highlight stronger communication between the cerebellum and other brain regions involved in language processing. Strengthened connections in these three regions could be associated with improved motor coordination and speech production, resulting in increased AQ and accompanying subscores.^34-35^ Furthermore, the cerebellum is also involved in higher order linguistic processes such as grammatical processing, lexical access, and verbal working memory – aspects commonly impaired in aphasia.^36-37^

The MCP is the major afferent pathway to the cerebellum - originating from the pontine nuclei, which integrate information from various cortical areas to establish the cortico-ponto-cerebellar system.^38^ Inputs from sensory, motor, and associative areas of the cerebral cortex are directed to the ipsilateral pontine nuclei, which in turn project to the contralateral cerebellar cortex through the MCP. Hence, the MCP is crucially involved in cognitive function and motor coordination.^39^ With higher FA in the MCP reflecting better structural white matter integrity, we speculate that in turn it would be associated with better neural communication between the cortex and cerebellum, leading to better language function.

The inferior cerebellar peduncle interfaces the spinal cord and medulla oblongata with the cerebellum, with afferent fibers passing lateral to the spinal tract of the trigeminal nerve.^40^ It receives proprioceptive information from the spinal cord and inputs from nuclei in the medulla, participating in the cerebellum’s role of balance and fine-tuning movements. As the left side of the cerebellum seems to be associated with timing and sequencing of precise motor planning, the L-ICP, which primarily conveys ipsilateral proprioceptive information from the spinal cord, likely plays a crucial role in supporting the temporal coordination of motor tasks such as speech production.^9-11,40^ This functional alignment may explain why the L-ICP demonstrates a stronger association with language function than the right ICP. We speculate that overall, higher structural white matter integrity in the L-ICP may correspond with enhanced connectivity between the aforementioned regions, leading to improved integration of proprioceptive information and movement, contributing to better language capabilities via smoothing of speech production.^40^

In our study, the FA of the region of the L-CST below the tentorium was found to be correlated with spontaneous speech and aphasia quotient. The corticospinal tract, part of the pyramidal tract, is one of the major neuronal pathways connecting the spinal cord and motor regions of the cortex.^41^ It plays a major role in cortical control of spinal cord activity and thus is the principal motor pathway for voluntary movements.^41^ Since motor nuclei needed for phonation and articulation are located from the pons down to the spinal cord’s lumbar portion, integrity of the corticospinal tract is heavily associated with speech production.^35^ The left hemisphere houses critical language areas such as Broca’s area, which formulates articulatory information to be executed predominantly by the left motor cortex.^22^ Given that the left CST originates ipsilaterally from the left motor cortex before crossing at the medulla, it serves as a critical pathway for transmitting motor commands essential for speech production to the spinal cord.^41^

This anatomical basis, coupled with the fact that our participants mainly had cortical lesions in the left perisylvian area, likely explains its stronger association with speech-related functions compared to the right CST. As higher FA indicates less damage to the structure of the white matter tracts of the infratentorial L-CST, we postulate a more robust pathway between cortical brain regions and the motor nuclei recruited in voluntary generation of speech.^41^

When considering the statistical significance of the left arcuate lesion volume’s association with WAB-R scores in our sample, it appears that the structural integrities of the MCP, L-ICP, and L-CST also show notable correlation with aphasia severity following stroke. Despite the arcuate fasciculus’ well-documented role in language processing and aphasia, drawing inferences from our data, FA of the infratentorial white matter tracts hold similar predictive potential.^23,42-44^ The statistical significance of univariate linear regression models using the MCP, L-ICP, and L-CST compared to that of the left arcuate lesion volume again suggests that these infratentorial brain regions can provide a substantial and consistent basis for examining WAB-R scores after a stroke.

These findings not only underscore the complexity of brain networks involved in language function, but highlight a critical insight: the neuroanatomical substrates of aphasia extend beyond conventional language circuits, possibly incorporating a wider range of mechanisms that include cerebellar and corticospinal involvement.^45-47^ The predictive power and correlational strength of infratentorial regions also provide a possible solution to the methodological challenges posed by extensive cortical damage in PSA patients. This focus on such regions could lead to better feasibility and ease of automation of evaluating PSA, especially when compared to the investigation of supratentorial areas. Future research should further explore these infratentorial white matter regions to develop more robust and clinically applicable biomarkers for PSA, potentially offering new avenues for diagnosis and treatment planning in cases where traditional supratentorial analyses are limited.

## LIMITATIONS

This study is subject to certain limitations. Even though the participants primarily experienced damage to the left superior longitudinal fasciculus, the differences in individual stroke lesions warrant a careful, holistic interpretation of results. This heterogeneity of stroke lesions among participants suggests the need for a larger sample size to better account for this variability. A larger cohort would allow for a more nuanced understanding of how different lesion characteristics, such as size and location, influence the relationship between white matter integrity and language outcomes.

However, given the significance of the correlations between white matter integrity and aphasia severity in infratentorial white matter regions including both cerebellar pathways and the corticospinal tract, we believe our findings provide valuable insights into the broader neural network involved in post-stroke aphasia. These results highlight the importance of considering both cerebellar and corticospinal pathways in understanding language deficits following stroke.

Another limitation of this study is the confined scope of our analyses to infratentorial white matter integrity (i.e., FA), excluding supratentorial white matter integrity from our investigation. This approach was necessitated by the prevalence of lesions in supratentorial areas, which often impede accurate delineation and quantification of white matter integrity in traditional language-associated regions. The relative preservation of infratentorial structures in PSA patients provided a unique opportunity to investigate potential biomarkers and neural correlates that are less confounded by stroke-induced structural changes, offering methodological advantages in terms of feasibility and reliability. However, this focus potentially overlooks the complex interplay between supratentorial and infratentorial networks in language processing. Even though our investigation of the relationship between the left arcuate fasciculus lesion volume and language function allows an exploration of the supratentorial region’s influence, without a direct analysis between the arcuate fasciculus and infratentorial regions we cannot completely mitigate this concern.

## CONCLUSION

Our study provides novel insights into the role of infratentorial white matter integrity, particularly in the middle cerebellar peduncle, left inferior cerebellar peduncle, and left corticospinal tract, in post-stroke aphasia severity. These regions demonstrated significant correlations with language performance and emerged as reliable predictors of aphasia severity, underscoring their potential as biomarkers. Importantly, by shifting focus to relatively preserved infratentorial structures, this approach circumvents challenges associated with cortical lesions, offering a more consistent framework for evaluating aphasia in patients with varying lesion profiles. The findings emphasize the involvement of cerebellar and corticospinal pathways in language processing, expanding traditional models that have predominantly focused on supratentorial regions. This expanded understanding may inform future clinical assessments and therapeutic interventions targeting not only cortical but also cerebellar and corticospinal pathways. Future research should explore the combined contributions of both infratentorial and supratentorial regions to offer a more comprehensive understanding of post-stroke aphasia mechanisms.

## Data Availability

The data related to this article will be shared upon reasonable request to the corresponding author.

## Funding

This work was funded by the Ability Central Foundations.

## Competing Interests

The authors declare that the research was conducted in the absence of any commercial or financial relationships that could be construed as a potential conflict of interest.

## Notes

### Competing Interest Statement

The authors have declared no competing interest.

### Author Declarations

IRB of Casa Colina Hospital and Centers for Healthcare gave ethical approval for this work.

